# Cannabis use, microbial diversity, and *Dialister* abundance in older adults with HIV: a cross-sectional study

**DOI:** 10.1101/2025.11.03.25339401

**Authors:** Donald D Porchia, Yan Wang, Zhi Zhou, Mingkai Chen, Eric C Porges, Ronald A Cohen, Smita Ghare, Shirish Barve, Robert L Cook, Zhigang Li

## Abstract

**Objectives:** People living with HIV (PLWH) frequently experience gastrointestinal symptoms linked to dysbiosis, impaired mucosal barrier integrity, and persistent immune activation. Cannabis is widely used for symptom management by PLWH, but its effects on the gut microbiome are unclear.

**Methods:** We conducted a cross-sectional analysis of 63 PLWH (mean age 59.4 years; 71.4% Black or Hispanic) enrolled in the Marijuana Associated Planning and Long-term Effects study and its microbiome substudy. Participants provided fecal samples for 16S rRNA sequencing. Cannabis use was quantified using a validated Timeline Followback. Alpha diversity was estimated using the Shannon index, beta diversity with Bray-Curtis dissimilarity and PERMANOVA, and genus-level abundance using the IFAA method. Models adjusted for sex, age, and education.

**Results:** Higher cannabis consumption was significantly associated with reduced alpha diversity (β = −1.23, *p*=0.038). No significant differences in beta diversity were observed between high and low-to-no groups (*p*=0.35). At the genus level, *Dialister* abundance showed a significant dose-dependent association with cannabis use, with a 14.4% reduction in abundance per 50 mg increase in THC per use-day (q=0.034). Reduced alpha diversity and *Dialister* depletion are notable given links to impaired mucosal barrier integrity, microbial translocation, and systemic immune activation in HIV.

**Conclusion:** Cannabis consumption in PLWH was associated with lower microbial diversity and reduced *Dialister* abundance, a taxon with dual roles in mucosal integrity and gastrointestinal symptom modulation. These findings suggest cannabis may modify HIV-associated dysbiosis, warranting further longitudinal studies to disentangle symptomatic benefits from long-term impacts on mucosal health and systemic inflammation.

## 1 Introduction

Antiretroviral therapy (ART) has significantly improved the prognosis of people living with HIV (PLWH). For individuals who initiated treatment after 2015 with CD4 counts ≥ 500 cells/µL, life expectancy approaches that of the general population^1^. Despite virologic suppression, HIV remains a chronic immune-mediated condition characterized by persistent inflammation, immune activation and multiple comorbid symptoms^2,3^. Gastrointestinal (GI) disturbances, such as abdominal pain, nausea, diarrhea and altered bowel habits, are common in PLWH and may be exacerbated by ART-related toxicities. Epidemiologic studies further indicate elevated risks for functional and inflammatory bowel diseases such as irritable bowel syndrome (IBS) and inflammatory bowel disease (IBD) compared with HIV-negative populations^4–7^.

The gut microbiome is integral to mucosal immune regulation, epithelial barrier integrity, and regulation of systemic inflammation^3,8,9^. In PLWH, gut dysbiosis, characterized by reduced alpha diversity, depletion of beneficial commensals, and enrichment of pro-inflammatory taxa, contributes to microbial translocation and chronic immune activation^3,8,9^, ultimately leading to systemic inflammation and immune dysregulation^3,8,10^. IBS and IBD, both more common in PLWH, are similarly associated with dysbiotic profiles that can exacerbate mucosal immune activation and barrier dysfunction^11,12^.

Cannabis use is common among PLWH, often for relief of HIV or ART-related symptoms, such as pain, anxiety, sleep disturbances and GI discomfort^13–15^. The bioactive cannabinoids in cannabis engage the endocannabinoid system (ECS), a signaling network with CB1 and CB2 receptors expressed in immune cells and throughout the gastrointestinal tract^16–19^. By modulating intestinal inflammation, barrier integrity, and microbial composition, the ECS provides a plausible pathway through which cannabis could influence HIV-related gut dysbiosis and downstream immune activation. Prior studies have linked problematic cannabis use, defined as having a Cannabis Use Disorder Identification Test (CUDIT)^20^ score ≥ 8, to alterations in gut microbial homeostasis in PLWH^21,22^, but the effects of general (non-problematic) cannabis use on microbial diversity and taxon abundance remain poorly understood.

Given the interplay between gut microbiota, mucosal immunity, and HIV pathogenesis, determining whether cannabis use modifies microbial diversity or specific taxa abundance may clarify mechanisms affecting GI comorbidities, systemic inflammation, and long-term disease outcomes in PLWH. In this cross-sectional analysis of the Marijuana Associated Planning and Long-term Effects (MAPLE) study and its microbiome substudy, we examine the association between cannabis use, gut microbial diversity, and genus-level absolute abundance in a cohort of older PLWH. We hypothesized that cannabis consumption would be associated with reduced alpha diversity and alterations in the gut microbiome composition.

## 2 Methods

### 2.1 Study Design

Data were drawn from the Marijuana Associated Planning and Long-term Effects (MAPLE) study and its supplemental substudy, which incorporated gut microbiome data from fecal samples and neurocognitive assessments. MAPLE is a longitudinal, prospective cohort study designed to examine the health effects of cannabis use among PLWH in Florida. The supplemental substudy focused specifically on cannabis use, the gut microbiome, and mild cognitive impairment (MCI) as a subsample of the MAPLE study. Data collection occurred between 2018 and 2022. Full details of the parent study have been published previously^23,24^.

### 2.2 Study Participants and Recruitment

Participants were recruited from community and public health clinics located in Alachua (Gainesville), Hillsborough (Tampa), and Miami-Dade (Miami) counties, Florida. All study procedures were approved by the Institutional Review Boards at the University of Florida, Florida International University and the Florida Department of Health. Written informed consent was obtained from all participants prior to enrollment.

The analytic sample included 63 participants (52.4% female, 71.4% Black or Hispanic, mean age: 59.4 years) who provided fecal samples for microbiome profiling. Two individuals were excluded as outliers based on their THC consumption per use-day using Grubbs’ test (THC > 1,000 mg per use-day) via the outlier package in R. Sociodemographic characteristics are presented in Table 1.

**Table 1.**
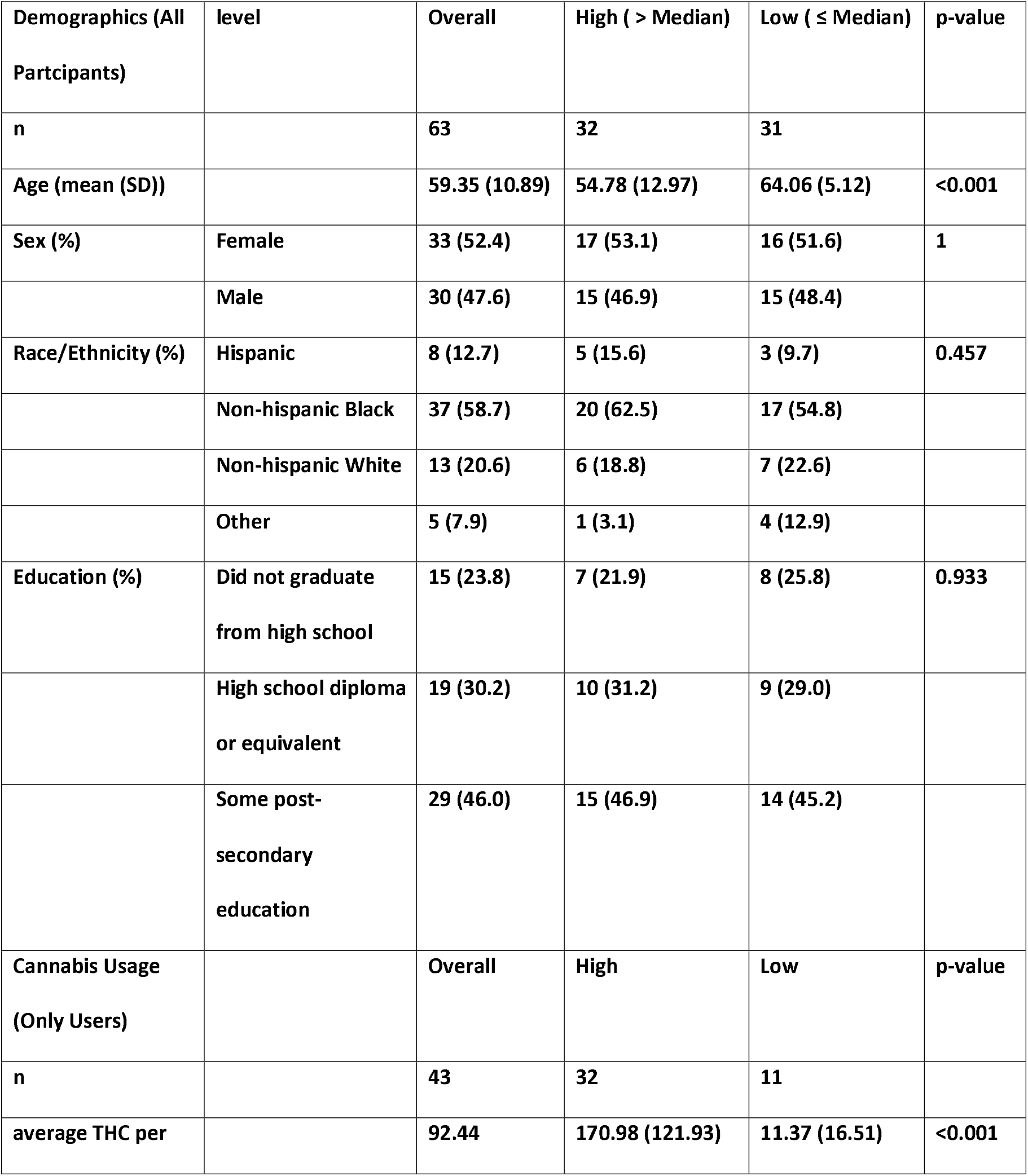
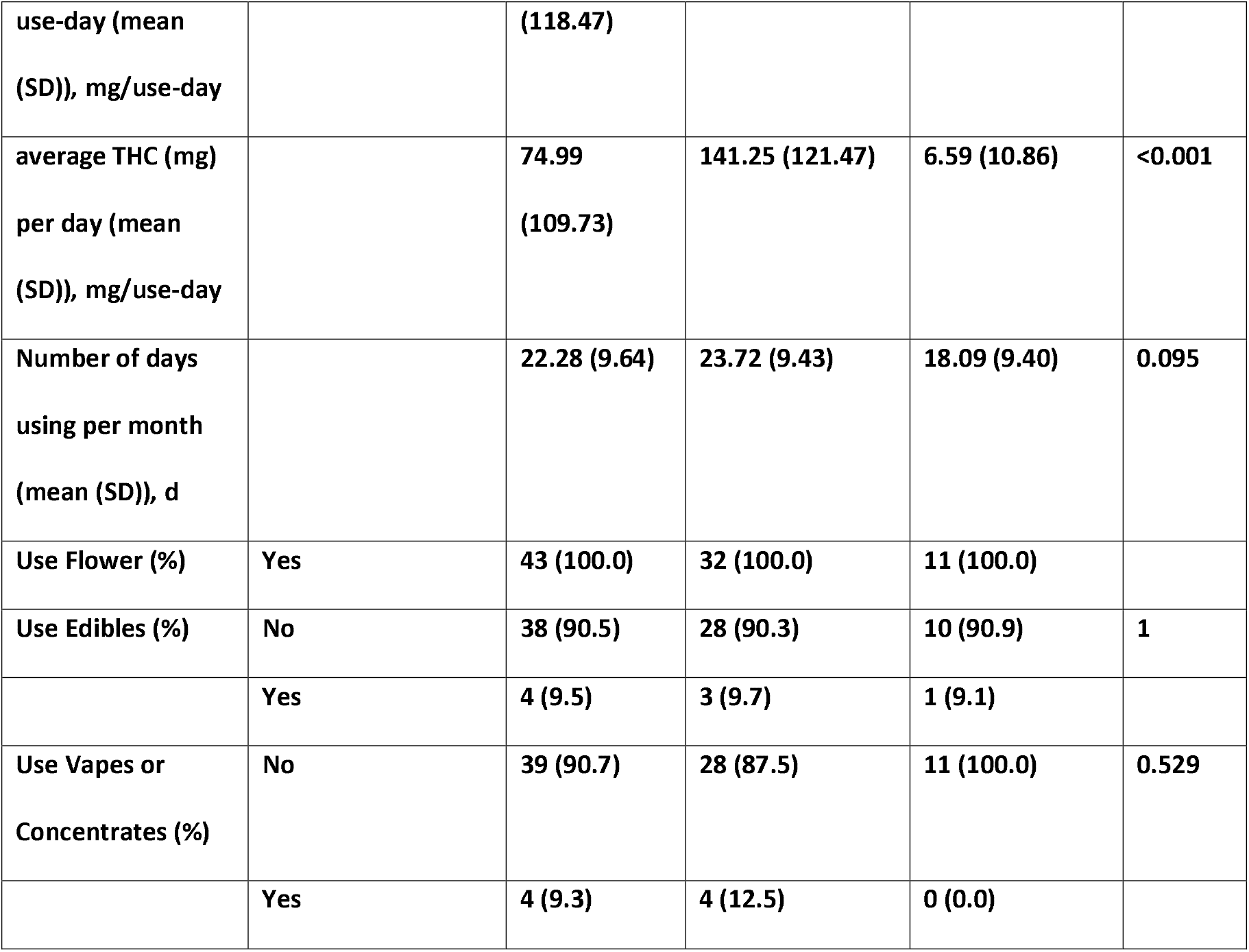
Sample sociodemographics.

### 2.3 Inclusion and Exclusion Criteria

The MAPLE study recruited both current cannabis users and non-users. Inclusion criteria were: (1) age ≥ 18, (2) confirmed HIV-positive status based on medical or prescription records, (3) intent to reside in Florida for the next 12 months, (4) English-speaking, and (5) verified current cannabis user or non-user status. Current cannabis users required a positive urine screen for THC and self-reported cannabis use ≥ 4 times/month. Non-users were defined by a negative urine screen for THC, no cannabis use in the past five years, and lifetime use ≤ 1 time/month.

Participants in the supplemental substudy were required to meet MAPLE eligibility criteria and additionally, be ≥ 60 years old with a clinical determination of mild cognitive impairment (MCI) or clinically confirmed absence of MCI.

### 2.4 Measure of cannabis use frequency and quantity

Cannabis consumption was assessed using a retrospective, calendar-based Timeline Followback (TLFB) adapted from an original version used in alcohol research. The TLFB has been previously validated for use with cannabis and other substances, such as cocaine and tobacco^25–27^. Trained MAPLE research assistants administered the TLFB to collect detailed data on cannabis use over the past 30 days preceding each study visit.

Participants reported frequency of use, method of consumption (e.g., joints, blunts, bowls, concentrates, or edibles), and the quantity of cannabis flower consumed in grams. When the THC concentration was known, flower-based consumption was converted to milligrams of THC; otherwise, a standard assumption of 15% THC per gram was applied. For cannabis concentrates, THC oil, and edibles, consumption was recorded directly in milligrams of THC per dose; see Table 2 for details. Non-users were coded as having zero cannabis consumption.

**Table 2.** THC per dosage of non-flower cannabis products.

| Non-Flower Cannabis Product | THC Concentration |
| --- | --- |
| THC oil | 4 mg per hit/puff/dose if given otherwise 10 mg per day |
| Edible (chocolate or butter) | 20 mg per dose |
| Edible (gummy) | 10 mg per dose |

### 2.5 Fecal sample collection and microbiota characterization by 16S rRNA gene sequencing

All participants were provided with a stool nucleic acid collection and preservation kit (Norgen Biotek Corp., Thorold, ON, Canada) with visual and printed instructions. Fecal samples were frozen at ≤ ™20°C until processing. The 16S rRNA gene sequencing methods were adapted from the published methods^28–30^. Briefly, total genomic DNA was extracted using the Qiagen MagAttract PowerSoil DNA Kit (formerly sold by MO BIO as PowerMag Soil DNA Isolation Kit). The 16S rRNA hypervariable region 4 (V4) was amplified by PCR using the barcoded Illumina adaptor-containing primers 515F and 806R and sequenced on the MiSeq platform (Illumina, San Diego, CA, USA) using the 2 × 250 bp paired-end protocol. The primers used for amplification contained adapters for MiSeq sequencing and a single-index barcode on the reverse primer so that the PCR products were pooled and sequenced directly, targeting at least 10,000 reads per sample. The read pairs were demultiplexed based on the unique molecular barcodes, and reads were merged using the USEARCH v7.0.1090. The 16S rRNA gene sequences were clustered into operational taxonomic units (OTUs) at a similarity cutoff value of 97% using the UPARSE algorithm^31,32^. The OTUs were mapped to the SILVA v128 database containing only the V4 region sequences to determine the taxonomies^33^. Abundances were recovered by mapping the demultiplexed reads to the UPARSE OTUs.

After data quality control and cleaning, 253 genera were detected across the population of 63 fecal samples collected. Each participant contributed one sample to the analyses in this cross-sectional study.

### 2.6 Statistical analysis

Statistical analyses of microbiome data were performed in R (version 4.3.3) using vegan (version 2.6-8)^34^ and the IFAA^35^ package for robust inference for absolute abundance.

Alpha diversity, a measure of within-sample microbial diversity, was quantified using the Shannon diversity index. Higher alpha diversity is generally considered indicative of a healthier gut microbiome. Multiple linear regression was used to assess the association between average THC consumed per use-day and the Shannon diversity index, adjusting for age, sex and education.

Beta diversity, reflecting between-sample differences in microbial composition, was assessed using Bray-Curtis dissimilarity. Permutational multivariate analysis of variance (PERMANOVA) was conducted with 1,000 permutations using the adonis2 function from the vegan package to compare gut microbiome composition between participants with high versus low-to-no use. High cannabis use was defined as an average THC consumption greater than the median of 56.25 mg per use-day; low-to-no use was defined as 56.25 mg or less, including non-users.

Genus-level associations with cannabis use were assessed using the IFAA method across 253 detected genera. All models were adjusted for age, sex and education. P-values were corrected for multiple testing using the Benjamini-Hochberg false-discovery rate (FDR) method^36^.

## 3 Results

### 3.1 Characteristics of study participants and cannabis consumption

Sociodemographic characteristics of the study population are shown in Table 1. The study population comprised older adults (mean age 59.4 years), the majority of whom were Black or Hispanic (71.4%) and female (52.4%). Participants had been living with HIV for an average of 20.4 years.

Cannabis use was reported by 70.5% of participants, with most obtaining cannabis outside of Florida’s medical marijuana program. Among the 43 cannabis users, 35 (81.4%) smoked cannabis flower exclusively, 4 (9.3%) used a combination of cannabis flower and edibles, and 4 (9.3%) used some combination of cannabis flower and either vapes or concentrates.

Cannabis users reported an average of 22.3 use-days in the past 30 days, with 41.9% reporting daily cannabis use. On average, participants consumed 75.0 (109.7) mg of THC per day and 92.4 (118.5) mg of THC per use-day. As there is no universally accepted standard THC dose analogous to a standard alcoholic drink^37–39^, we categorized participants into high and low consumption groups based on the sample median of 56.25 mg per use-day. Among the 63 participants, 32 were classified as high consumers and 31 were low-to-no consumers (11 users plus 20 non-users). Notably, only participants in the high consumption group reported using vapes or concentrates.

### 3.2 Alpha diversity

The Shannon diversity index reflects both the richness and evenness of microbial taxa within a sample population with higher values indicating greater diversity and a more uniform distribution. In our regression analysis, we found a statistically significant negative association between cannabis consumption and Shannon diversity (β = −1.23, *p* = 0.038), suggesting that higher cannabis use is associated with reduced within-sample microbial diversity. This association is visualized in Figure 1.

**Figure 1.**
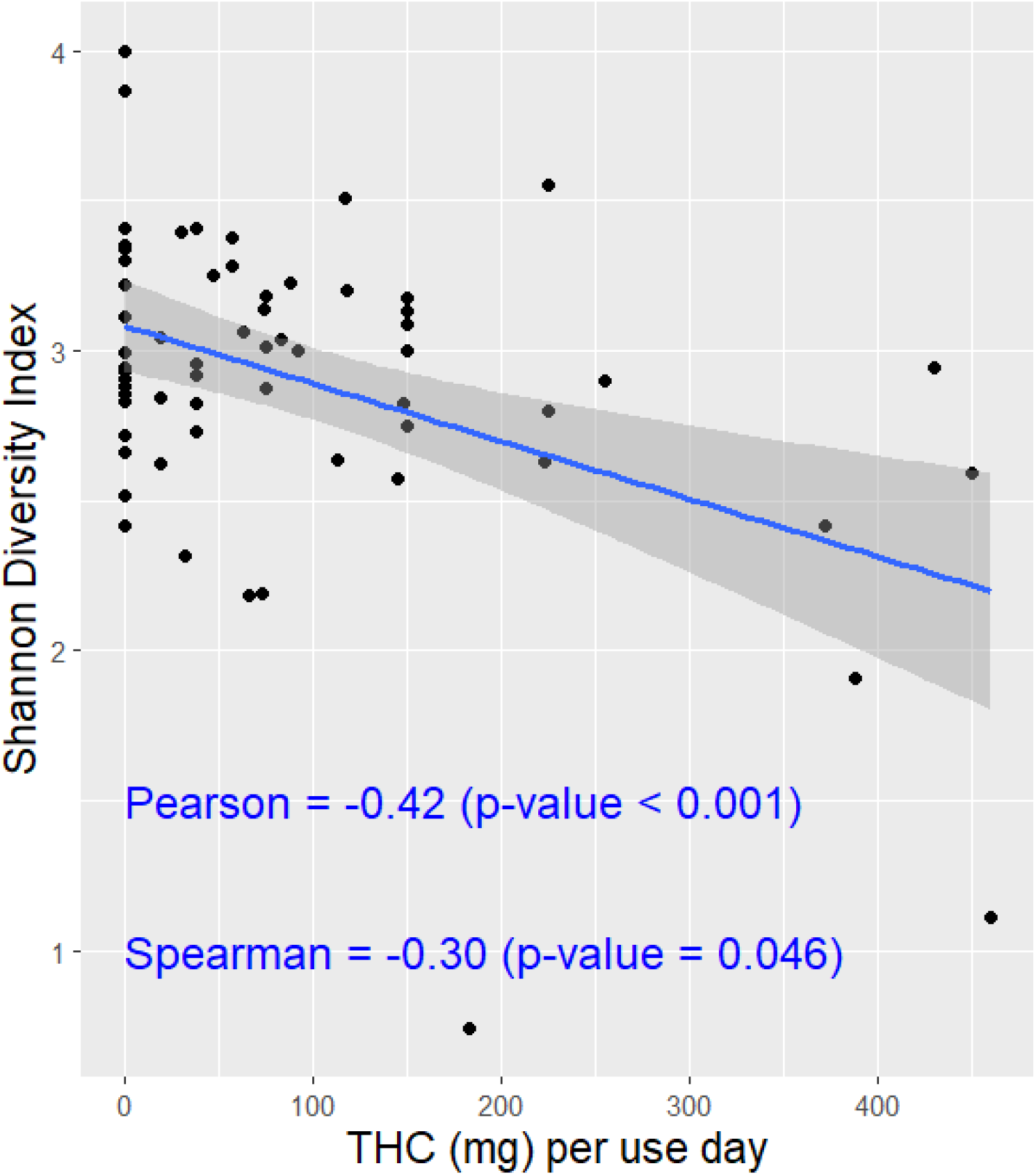
Association between cannabis use and gut microbial alpha diversity among people living with HIV. The plot displays the relationship between the gut microbiome diversity, measured by the Shannon index and cannabis use, quantified as milligrams of THC per use-day. A statistically significant negative association suggests reduced microbial diversity with increasing cannabis use. Results were adjusted for sex, age and education.

### 3.3 Beta diversity

Beta diversity measures the between-subject variation in gut microbiome composition. In this study, we compared beta diversity between participants categorized into high versus low-to-no cannabis consumption groups, using a median split of 56.25 mg of THC per use-day.

Bray-Curtis dissimilarity was used to assess compositional differences, and results were visualized via Principal Coordinates Analysis (PCoA) as described in Section 2.6. Although the ordination plot (Figure 2) shows some visual separation between groups, there was substantial overlap. PERMANOVA analysis with 1,000 permutations, adjusted for age, sex and education, indicated no statistically significant difference in overall gut microbial composition between the high and low-to-no use groups (*p* = 0.35). These results suggest that cannabis consumption was not a major factor explaining between-sample variation in microbiome composition in this cohort.

**Figure 2.**
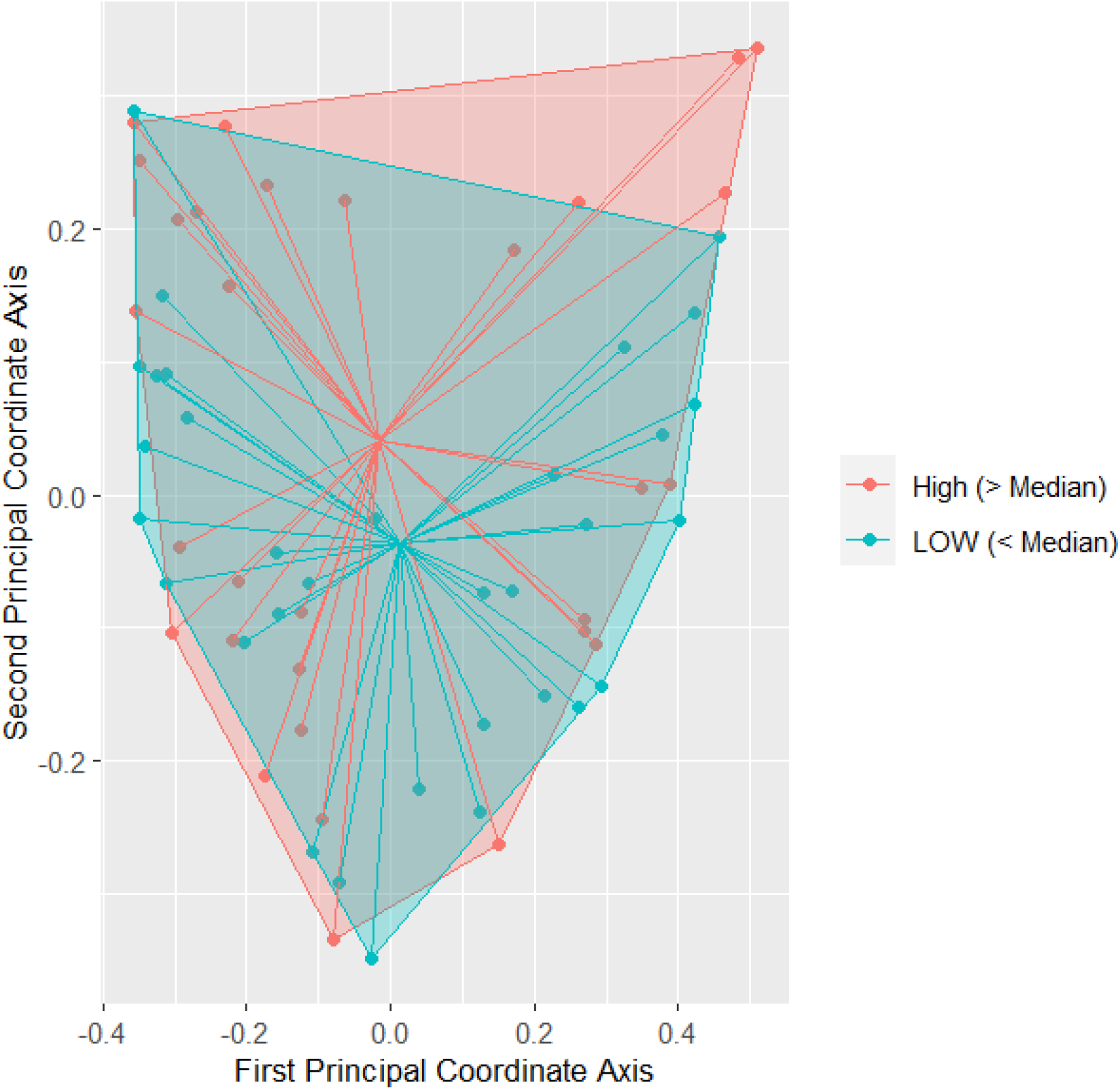
Beta diversity of gut microbiome composition among high and low-to-no cannabis use groups in people living with HIV. Principal Coordinates Analysis (PCoA) of Bray-Curtis dissimilarity from fecal 16S rRNA profiles. Each point in an individual sample colored by use group (high > 56.25 mg THC per use-day, n = 32; low-to-no ≤56.25 mg THC per use-day, n = 31). While some visual separation is present, the groups show substantial overlap. Differences in overall community composition were not statistically significant by PERMANOVA using 1,000 permutations and adjusting for age, sex and education (*p*=0.350).

### 3.4 Genus-level absolute abundance

We analyzed the absolute abundance (AA) of microbial genera from the phylogenetic tree in relation to cannabis consumption using the IFAA method. AA represents the quantity of bacteria present in the gut microbiome. Cannabis use was modeled as average THC consumption per use-day, and all models were adjusted for age, sex and education. P-values were corrected for multiple comparisons using the FDR method.

Of the 253 detected genera, only *Dialister* showed a statistically significant association with cannabis use after FDR correction. Specifically, the AA of *Dialister* decreased by 14.4% per 50 mg increase in THC per use-day (*q* = 0.034), suggesting a dose-dependent negative relationship. This finding indicates that high cannabis consumption may reduce the presence of *Dialister* in the gut ecosystem. A heatmap of all genus-level associations with cannabis use is presented in Figure 3, with color gradients representing the direction and magnitude of the associations.

**Figure 3.**
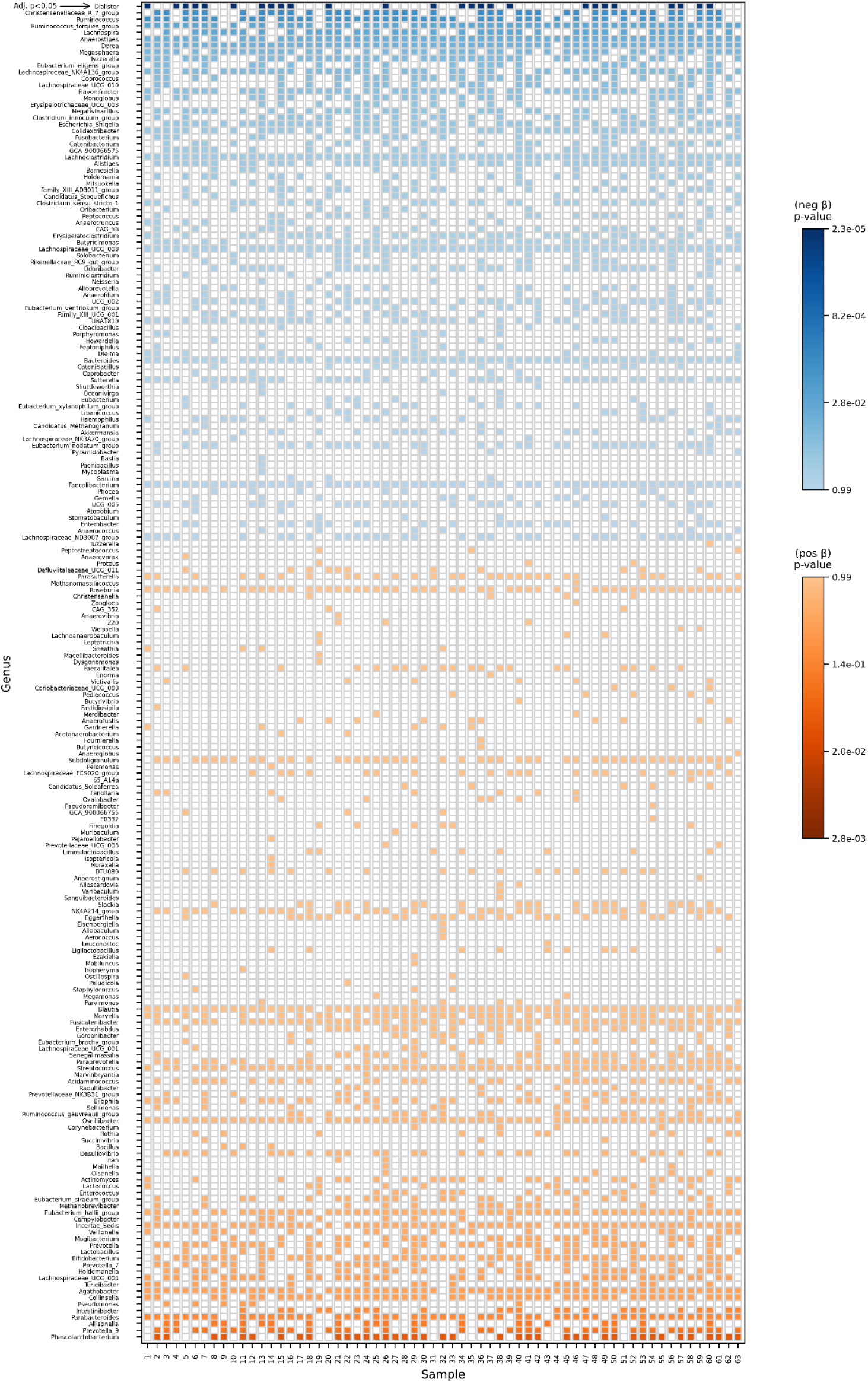
Genus-level associations between cannabis use and absolute abundance in people living with HIV. Heatmap of IFAA-derived significance and direction of associations between genera and cannabis use measured in milligrams of THC per use-day. Blue indicates negative associations, while orange indicates positive associations. The genus *Dialister* (at the top) showed a statistically significant negative association after adjusting for multiple testing and controlling for sex, age and education.

## 4 Discussion

In this study of older PLWH, we observed that higher cannabis consumption was associated with reduced alpha diversity and decreased absolute abundance of the genus *Dialister*. These findings add to the limited literature on cannabis-microbiome interactions and provide novel insights into how cannabis may shape gut microbial ecology in the context of HIV.

Reduced alpha diversity is a well-recognized feature of HIV-associated dysbiosis and has been directly linked to impaired mucosal barrier integrity, microbial translocation, and chronic systemic immune activation^2,3,8,10^. In HIV, disruption of the epithelial barrier allows bacterial products such as lipopolysaccharide (LPS) to enter systemic circulation, perpetuating inflammation and fueling non-HIV comorbidities^2,8,10^. Our observation that greater cannabis use correlated with lower Shannon diversity suggests that cannabis may further compromise microbial stability in a population already vulnerable to barrier dysfunction.

The genus *Dialister* showed a dose-dependent negative association with cannabis exposure, decreasing in abundance by an average of 14.4% per 50 mg increase in THC per use-day. This result has dual implications. On one hand, reductions in *Dialister* may exacerbate HIV-related mucosal damage, given prior evidence linking fluctuations in *Dialister* abundance to altered short-chain fatty acid metabolism, mucosal inflammation, and epithelial permeability^11,12,40^. In this framework, cannabis-driven *Dialister* depletion could worsen microbial translocation and amplify immune activation, processes that remain central to HIV pathogenesis even under ART^2,8,10–12,40^.

On the other hand, studies in non-HIV populations have reported that decreases in *Dialister* can coincide with improved GI symptoms. For example, patients with IBS who responded to fecal microbiota transplantation (FMT) experienced both symptom relief and reductions in Dialister^41–43^. Similarly, for IBD, higher levels of *Dialister* have been linked to microbial dysbiosis, increased mucosal inflammation and unfavorable clinical outcomes, indicating that alterations in this genus may be tied to disease severity^12,40,44^. These observations suggest that high *Dialister* abundance may also contribute to dysregulated mucosal immune signaling in some contexts. From this perspective, cannabis-related reductions in *Dialister* could partially explain why many PLWH report subjective improvement in GI discomfort when using cannabis.

Taken together, our findings highlight the complexity of cannabis-microbiome interactions. Cannabis consumption may provide symptomatic relief through modulation of taxa such as *Dialister*, while at the same time promoting features of dysbiosis, lower diversity and potential barrier impairment, that could intensify systemic immune activation in PLWH. These seemingly contradictory effects underscore the nonlinear nature of cannabis-microbiome interactions and the need for mechanistic studies that distinguish between short-term symptom benefits and long-term consequences for mucosal integrity and immune health.

### 4.1 Limitations

This study has several limitations. Recruitment from three Florida counties may limit generalizability, and the moderate sample size restricts the power to detect more subtle associations. Although the models were adjusted for sociodemographic covariates, unmeasured confounding may still exist. Most importantly, the cross-sectional design precludes causal inference, and temporal relationships between cannabis use and microbiome alterations cannot be established.

## 5 Conclusion

Cannabis use in PLWH was associated with reduced microbial alpha diversity and dose-dependent depletion of *Dialister*. These alterations have important implications for mucosal barrier integrity, microbial translocation, and systemic immune activation, which remain central to HIV pathogenesis despite effective ART. At the same time, evidence from non-HIV populations suggest that a reduction in *Dialister* has been associated with symptomatic relief in IBS and IBD, raising the possibility that cannabis provides short-term gastrointestinal benefit while also promoting long-term features of dysbiosis.

Overall, our findings highlight the complex and context-dependent nature of cannabis-microbiome interactions. Longitudinal and interventional studies are needed to determine whether cannabis consumption ultimately mitigates or exacerbates HIV-associated dysbiosis, and to disentangle symptom-level improvements from potential adverse consequences for immune health.

## Institutional Review Board Statement

All research procedures were approved by the Institutional Review Boards at the University of Florida, Florida International University, and the Florida Department of Health. Approval date: December 2017.

## Informed Consent Statement

Informed consent was obtained from all subjects involved in the study.

## Data Availability Statement

The data underlying this study are available from the corresponding author upon reasonable request. Requests should be submitted via the SHARC concept submission process (https://sharc-research.org/get-involved/submit-a-concept/) and will be provided subject to institutional approvals and a data use agreement.

## Author Contributions

Conceptualization: RLC, YW, ZL, DDP; Data Curation: ZZ; Formal analysis: DDP, ZL; Funding acquisition: RLC, YW; Data acquisition and Methodology: RLC, YW, ECP, SG, SB; Project administration: RLC, YW; Visualization: DDP, MC; Writing – original draft: DDP, ZL; Writing – review and editing: RLC, YW, ZL, DDP, ECP, RAC, SG, SB, ZZ, MC. All authors approved the final manuscript and agree to be accountable for all aspects of the work.

## Additional Contributions

We would like to thank all the participants and study staff who provided their time to make the Marijuana Associated Planning and Long-term Effects (MAPLE) study possible.

We would also like to thank the following list of organizations for their contributions: Care Resource Community Health Center, Hillsborough County Department of Health, Alachua County Department of Health, University of Miami Center for AIDS Research (CFAR), and University of Florida Infectious Disease Clinic.

